# Characterization of the humoral immune response to BNT162b2 in elderly residents of long-term care facilities five to seven months after vaccination

**DOI:** 10.1101/2021.11.09.21266110

**Authors:** Marla Delbrück, Sebastian Hoehl, Tuna Toptan, Barbara Schenk, Katharina Grikscheit, Melinda Metzler, Eva Herrmann, Sandra Ciesek

**Affiliations:** Institute of Medical Virology, Goethe University Frankfurt, Germany; Institute of Biostatistics and Mathematical Modeling, Goethe University Frankfurt; Fraunhofer Institute for Molecular Biology and Applied Ecology (IME), Branch Translational Medicine and Pharmacology, Frankfurt am Main, Germany; German Center for Infection Research, DZIF, Braunschweig, Germany

## Abstract

The elderly residing in long-term care facilities (LTCFs) are a group at high risk for COVID-19. Hence, monitoring of the vaccine-based immunity has a pivotal role in identifying strategies to provide optimal protection in this population. We examined the immune response to the mRNA vaccine BNT162b2 against COVID-19 five to seven months after completing a two-dose regimen.

We determined significantly lower anti-SARS-CoV-2 antibody titers in 298 SARS-CoV-2 naïve residents who were at least 75 years of age (mean 51.60 BAU/ml) (median age 87 years, range 75 to 101 years) when compared to health care workers (HCWs) aged 18 to 70 years (mean 156.99 BAU/ml, p < 0.001). Of the SARS-CoV-2 naïve residents, 29 had detectable neutralizing antibodies against the Delta variant (9.5%), and 14 of those (48.3%) only had a borderline titer of 1:10. Of 114 HCWs, 36 (31.6%) had detectable neutralizing antibodies. In a group of 14 elderly residents who had had a PCR-confirmed breakthrough infection, the mean antibody titer was significantly higher than in the other two groups (3199.65 BAU/mL) (p < 0.001), and 12 (85.7%) had detectable neutralizing antibodies against the Delta variant.

Our data demonstrate that 90.5% of elderly residents of LTCFs had no detectable neutralization-competent antibodies against the dominant Delta variant five to seven months after vaccination, and that neutralizing antibody titers were restored following a break-through infection. Our results suggest that both residents and health care workers in LTCFs would benefit from a booster vaccine six months after completing the two-dose schedule or earlier.

## Introduction

Advanced age is a strong risk factor for severe and fatal disease when infected with SARS-CoV-2.^1–3^ In the elderly, in contrast to children and younger adults, coronavirus disease of 2019 (COVID-19) is rarely asymptomatic^4^, and respiratory failure and organ dysfunction are present in many hospitalized patients.^5,6^ Therefore older adults were designated a high priority to be vaccinated in the roll-out of COVID-19 vaccines.^7^

Due to increased exposure to the virus, living in long-term care facilities (LTCFs) further increased COVID-19-related mortality in this group of patients^8^. In Europe, 30 to 60% of all COVID-19-related deaths were attributed to residents of LTCFs during the early pandemic.^9^ Early COVID-19 vaccine studies found a high efficacy in adults^10,11^, but the elderly, especially with comorbidities and frailty, were commonly underrepresented or excluded.^12^

Waning immunity due to immunosenescence can lead to impaired immunity in these high-risk patients.13 Vaccine-induced antibodies in the elderly display lower protective capacity, and T-cell response is skewed towards short-lived effectors.14 In case of COVID-19, the vaccine is usually given to establish novel immunity rather than boosting pre-existing immunity, which is the case in most vaccines applied in the elderly. Although first real-world COVID-19 vaccine efficacy data have been favorable, efficacy against hospitalization and death was lower in those 80 years of age and older when compared to 65 to 79 years of age.^15^

The emergence of novel variants of concern, most recently the Delta variant, and the concerns about waning immunity against symptomatic and severe COVID-19 raised a discussion on the need for a booster shot. But there is uncertainty regarding the effect of age and frailty on immunity against severe COVID-19 after vaccination.

In this study, we sought to determine whether markers of humoral and cellular immunity differ significantly between older adults (≥75 years of age), and a control group of health care workers (HCWs) six months after vaccination and analyzed the association of multiple variants, including medication and multimorbidity, on the immune response.

## Material and Methods

### Recruitment of study participants and inclusion criteria

This is a non-interventional observatory study. The Hessian Ministry for Social Affairs and Integration commissioned this study and approached long-term care providers in Hesse with suitable characteristics to participate in the study. All participants were informed about the aim of the study, and written informed consent was obtained from either the study participants themselves or from the legal guardian in case one had been appointed. Blood samples and data were collected at the LTC facilities. Three groups of study participants were formed.

The complete two-dose schedule of BNT162b2 (Comirnaty, BioNTech/Pfizer), applied at the recommended time interval of 21 days, had to be completed five to seven months before blood collection regardless of the study group. Further inclusion criteria for study participants in the main study group (*group 1*) of SARS-CoV-2 naïve residents in LTCFs for the elderly included age of at least 75 years at the day of their first vaccination. All participants with a known and confirmed SARS-CoV-2 infection in the past or a positive anti-SARS-CoV-2 nucleocapsid antibody test were excluded from the main analysis. In addition, two control groups were also recruited: HCWs in LTCFs for the elderly between the ages of 18 and 70 years were recruited to the first control group (*group 2*). HCWs with a known SARS-CoV-2 infection in the past or a positive anti-SARS-CoV-2 nucleocapsid antibody titer were excluded. The second control group (*group 3*) was comprised of residents of the age of 75 years or older with a subsequent, PCR-confirmed SARS-CoV-2 breakthrough infection no earlier than 14 days after the second vaccination.

### Markers of the humoral immune response

#### Anti-SARS-CoV-2 Nucleocapsid Antibody Assay

To determine whether a previous infection with SARS-CoV-2 had occurred, serum samples were tested for the presence of anti-SARS-CoV-2 nucleocapsid antibodies. We used the Abbott ARCHITECT SARS-CoV-2 IgG test (Abbott Laboratories. Abbott Park, Illinois, USA). Detected antibodies may be elicited after infection, but not vaccination with an mRNA vaccine.

#### Anti-SARS-CoV-2 Spike IgG Antibody Assay

Serum samples were tested for the presence of anti-SARS-CoV-2 Spike IgG antibodies. For this, we used the AdviseDx SARS-CoV-2 IgG II assay on the Abbott Alinity i® platform (Abbott Laboratories, Abbott Park, Illinois, USA). This assay detects antibodies targeted specifically against the receptor-binding domain of SARS-CoV-2. The results are provided in standardized binding antibody units (BAU) per ml. A result of less than 7.1 BAU/ml was considered negative, and a result of 7.1 to 8.51 BAU/ml was considered positive. Detectable antibodies may be elicited after both infection and vaccination with an mRNA vaccine.

#### Neutralization assay against the Delta variant

Serum samples were further analyzed for the presence of antibodies with neutralizing capacity against the Delta variant of SARS-CoV-2 (B.1.617.2) in a biosafety level 3 laboratory. The methodology of the SARS-CoV-2 neutralization assay has been described elsewhere.^16^

In brief, serum samples were serially diluted (1:2) and incubated with 4000 TCID50/mL of the Delta variant of SARS-CoV-2 (B.1.617.2) for one hour prior to infection of CaCo-2 cells. After 48 hours inoculation infected cells were examined for cytopathic effect (CPE) formation by light microcopy to determine the neutralization titer.

### Clinical parameters and patient history

Items were recorded in an interview with each study participant and derived from the patient history form with consent of the patient, or legal guardian if one had been appointed. Study participants from the HCW-group provided information on the items themselves. These items were: (i) date of birth (ii) dates of vaccination with BNT162B2 (iii) current height and weight (iv) known medical diagnoses (v) current medication.

### Statistical analysis

Data analysis was performed using RStudio Version 1.4.1717. Mean Anti-SARS-CoV-2 spike IgG antibody titers were tested for normality (Shapiro-Wilk test), and homogeneity of variance between the three study groups respectively (Levene’s test) The threshold for statistical significance was α < 0.05 in two-sided t-tests.

### Funding source and ethical approval

The study was commissioned and funded by the Hessian Ministry of Social Issues and Integration.The study protocol has been approved by the ethics board of the University Hospital Frankfurt (No. 20-864) and has been registered on the German Clinical Trial Register (DRKS00025813).

## Results

### Study population

Samples were collected from 22^nd^ July to 16^th^ September 2021 in 16 LTCFs in Hesse, Germany.

#### Group 1: Residents of LTCFs ≥ 75 years of age without prior infection or breakthrough infection

A total of 298 residents of the LTCFs participated in the study and were included in group 1. 212 (71.1%) were female, 86 (28.9%) were male, and none reported non-binary gender. The median age was 86 years (range: 75 to 101 years, IQR: 82 to 90.8 years). The mean Body Mass Index (BMI) was 25.6 kg/m^2^ ranging from 14.9 kg/m^2^ to 42.9 kg/m^2^.

#### Group 2: HCWs aged 18 to 70 years of age

114 HCWs were included in group 2. 83 (77.8%) were female, and 31 (27.2%) were male. None reported non-binary gender. The median age was 53 years (range: 24 to 70 years, IQR: 45.3 to 59.8 years). The mean BMI was 26.8 kg/m^2^ ranging from 18 to 50 kg/m^2^.

#### Group 3: Residents of LTCFs ≥ 75 years of age with breakthrough infection

14 residents of the LTCFs were included in group 3. 13 were female (92.9%), one was male (7.14%), none reported non-binary gender. The mean number of days between the second vaccination and the positive PCR test result was 109 days. The earliest breakthrough infection happened 19 days, and the latest 211 days after the second vaccination. The median age was 89 years (range: 82 to 93 years, IQR: 86.3 - 91). The mean BMI was 24.24 kg/m^2^ ranging from 17 to 32 kg/m^2^.

**Table 1** provides an overview of the study participants in all three groups.

**Table 1:** Characteristics of the study participants

|  | Participants<br>(n = 426) |
| --- | --- |
| **Role** | **[% of all participants]** |
| Group 1: *Residents ≥ 75 years of age* | 298 (70%) |
| Group 2: *HCWs aged 18 to 70 years of age* | 114 (26.8%) |
| Group 3: *Residents with breakthrough infection* | 14 (3.2%) |
| **Age at day of first vaccination** | **[median (IQR)]** |
| Group 1 | 86.0 (82.0 - 90.75) |
| Group 2 | 53.0 (45.25 - 59.75) |
| Group 3 | 89.0 (86.25 – 91) |
| **Sex** | **[% in each group]** |
| Group 1 |  |
| Female | 212 (71.1%) |
| Male | 86 (28.9%) |
| Group 2 |  |
| Female | 83 (72.8%) |
| Male | 31 (27.2%) |
| Group 3 |  |
| Female | 13 (92.9%) |
| Male | 1 (7.1%) |
| **BMI** | **[median in kg/m<sup>2</sup> (IQR)]** |
| Group 1 | 25.3 (22.4 - 28.6) |
| Group 2 | 26.4 (22.5 - 30.2) |
| Group 3 | 23.34 (20.47 - 28.64) |
| **Interval between second vaccination and blood draw** | 6.49 (5.08 – 7.70) [median in months (range)] |

### Markers of the humoral immune response

Of the study participants in group 1 (residents ≥ 75 years of age without known prior infection or breakthrough infection), 6 (0.02%) had detectable SARS-CoV-2 nucleocapsid antibodies and were excluded from the primary analysis. In group 2 (HCWS aged 18 to 70 years without a prior infection or breakthrough infection) no participant had detectable SARS-CoV-2 nucleocapsid antibodies. In group 3, which was comprised of residents with a PCR-confirmed breakthrough infection, only 4 of 14 participants (28.6%) had detectable anti-SARS-CoV-2 nucleocapsid antibodies.

50 participants (16.78%) in group 1 tested negative for anti-SARS-CoV-2 spike IgG antibodies, 9 (3.02%) had a borderline positive result and 239 (80.2%) tested positive. In the first control group of HCWs (group 2), a vast majority of 97.37% (111/114) of participants tested positive. Two participants tested negative, and one tested borderline positive. In the second control group of residents with a breakthrough infection (group 3), all 14 participants tested positive for anti-SARS-CoV-2 spike IgG antibodies (14/14) (**figure 1**).

**Figure 1:**
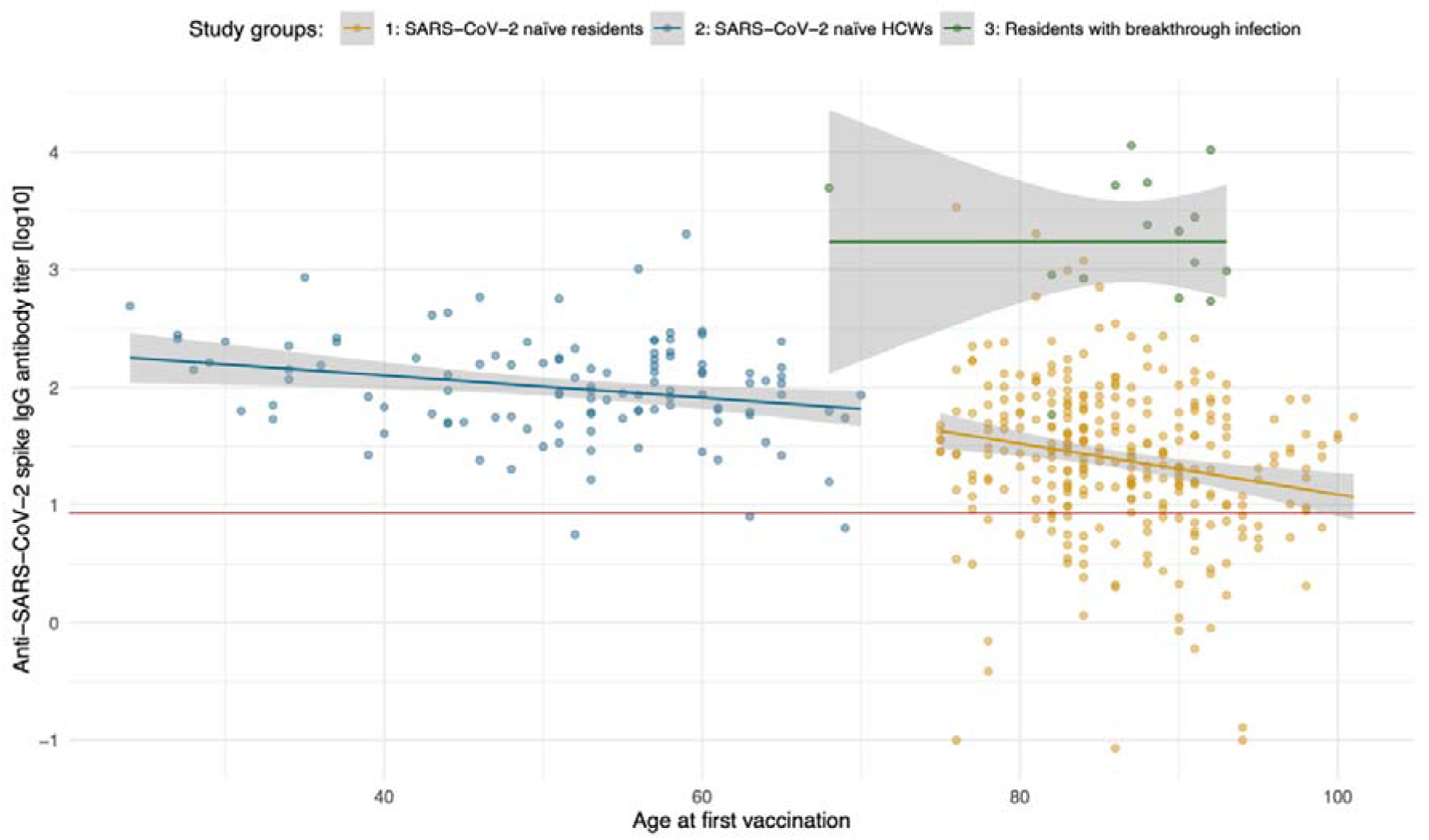
Anti-SARS-CoV-2 spike IgG titer (logarithmic) by study participants’ age. Group 1 (residents of LTCFs ≥ 75 years of age; orange), group 2 (HCWs at LTCFs, 18 to 70 years of age; blue), group 3 (residents of LTCFs after a breakthrough infection, green). Plotted within the scatter is the linear regression (lines in colors of the respective study groups) with the standard error (grey areas, 95% CI). The red line displays the cut-off anti-SARS-CoV-2 spike IgG antibody concentration of 8.52 BAU/ml, which is considered a positive test result (borderline: 7.10 - 8.51 BAU/ml).

In group 1, the anti-SARS-CoV-2 spike IgG antibody concentration had a mean of 51.60 BAU/ml (range: 0.90 – 710.64 BAU/ml, IQR: 12.09 – 61.47 BAU/ml). This was lower than in group 2, which had a mean of 156.99 BAU/ml (range: 5.61 – 2008.16 BAU/ml, IQR: 57.04 – 176.29 BAU/ml). This difference was statistically significant (CI 95%, p < 0.001) (**figures 1-3**). In these groups, a linear decline with age was observed, which was more prominent in group 1 (CI 95%, p < 0.001) (**figure 1**). In group 3, the mean antibody concentration of anti-SARS-CoV-2 RBD IgG was highest, with 3199.65 BAU/mL (range: 58.73 – 11360.00 BAU/ml, IQR: 857.65 – 4601.88 BAU/ml). This is significantly higher than in group 1 and group 2 (CI 95%, p < 0.001, respectively).

Neutralizing antibodies against the Delta variant were detected in 29 study participants (9.7%) of group 1, of which 14 (48.3%) had a borderline positive titer of 1:10. In group 2, 36 participants (31.6%) had detectable neutralizing antibodies against the Delta variant, of which 16 were borderline positive (titer of 1:10). In group 3, residents with a breakthrough infection, 12 (85.7%) had detectable neutralizing antibodies against the Delta variant, of which none were borderline (**figure 4**).

## Discussion

Here, we report the analysis of markers of the humoral response five to seven months after vaccination with the mRNA vaccine BNT162b2 in a group of residents of LTCFs. The SARS-CoV-2 naïve residents, aged ≥ 75 years of age, had a significantly lower concentration of anti-RBD-antibodies than younger health HCWs (**figures 2 and 3**). A linear decline with age was observed in both the group of residents and HCWs but was more marked in the elderly (**figure 1**).

**Figure 2:**
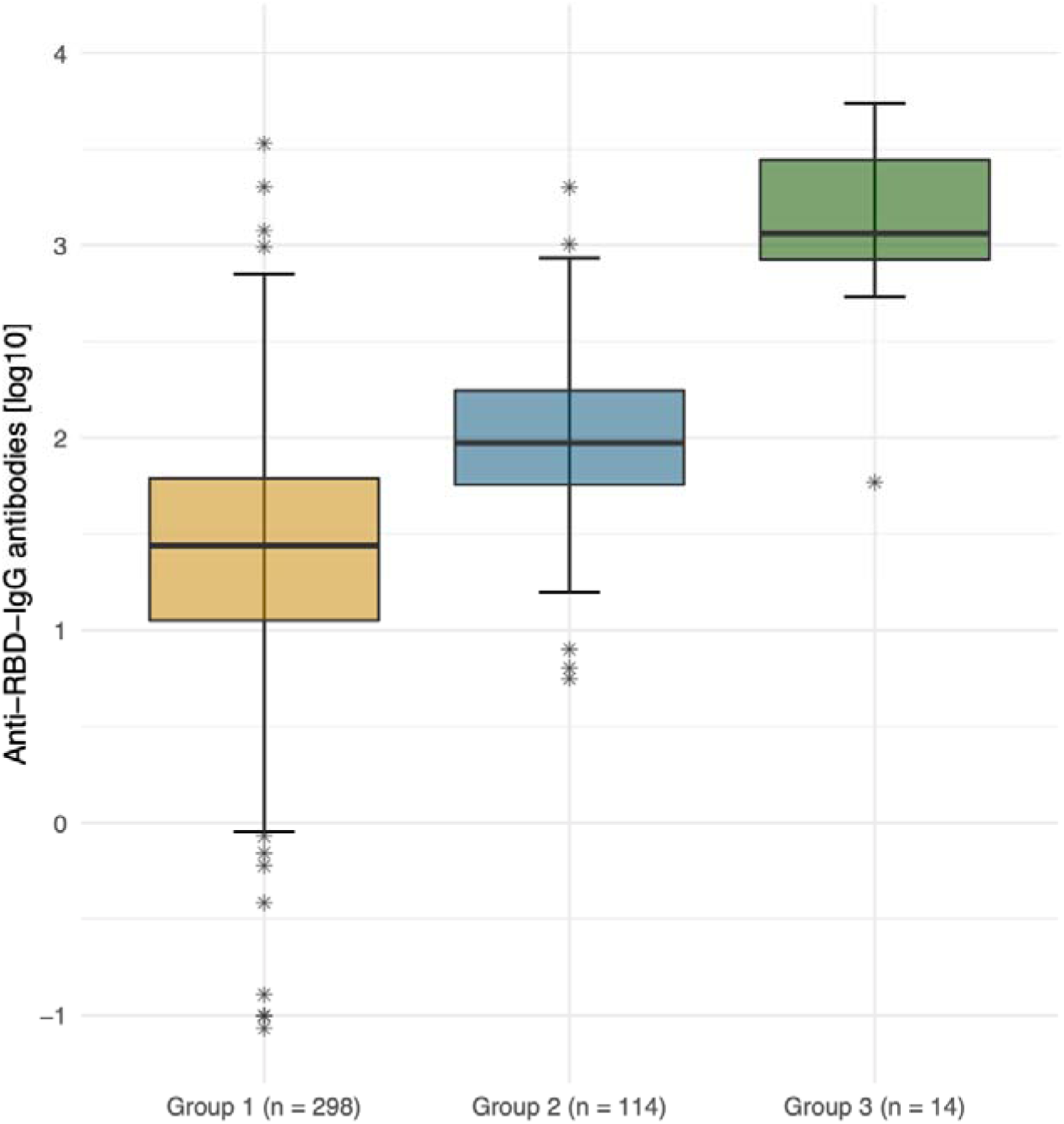
Anti-SARS-CoV-2 spike IgG antibody titers (logarithmic) by group. Logarithmic depiction in boxplots, 95% CI and IQR (25%-75%) of group 1 (residents of LTCFs ≥ 75 years of age; orange), group 2 (HCWs at LTCFs, 18 to 70 years of age; blue), and group 3 (residents of LTCFs after breakthrough infection, green). Stars represent outliers.

**Figure 3:**
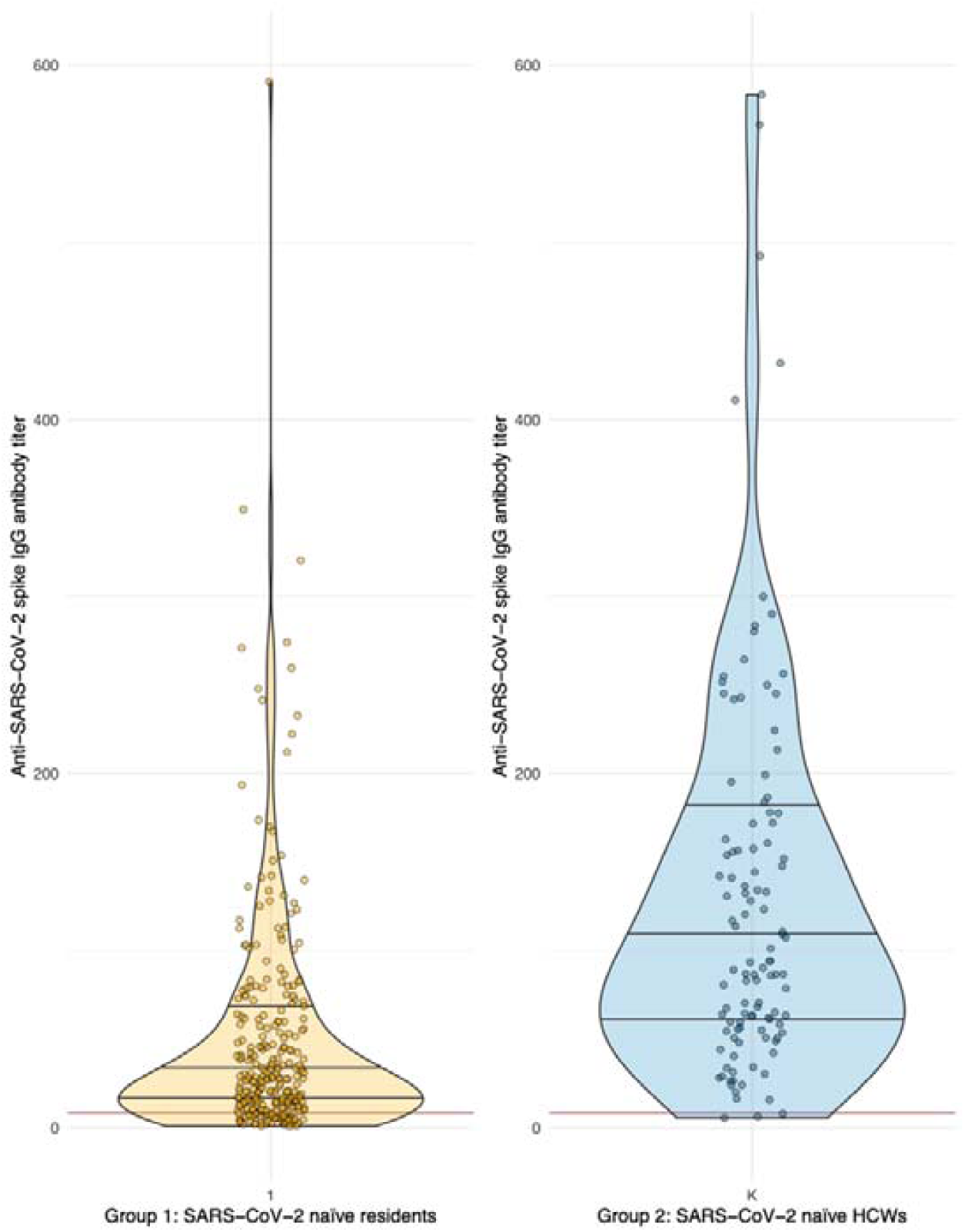
Anti-SARS-CoV-2 spike IgG antibody titer (non-logarithmic). Violin plots for visualization of differences in the distribution of anti-SARS-CoV-2 spike IgG antibody titers of groups 1 (residents of LTCFs ≥ 75 years of age; orange) and 2 (HCWs at LTCFs, 18 to 70 years of age; blue). The red line displays the cut-off anti-SARS-CoV-2 spike IgG antibody concentration of 8.52 BAU/ml, which is considered a positive test result (borderline: 7.10 - 8.51 BAU/ml).

The elderly also had a smaller fraction of individuals with detectable neutralizing antibodies against the Delta variant (9.7%, **figure 4**). Almost half (48.3%) of this minority with detectable neutralization only exhibited a borderline positive titer of 1:10. While the mean anti-SARS-CoV-2 IgG antibody concentration in our control group of younger HCWs was significantly higher, neutralization against the Delta variant was also only detected in a minority of 31.6% of the participants in this group five to seven months after having completed a full series of vaccination.

**Figure 4:**
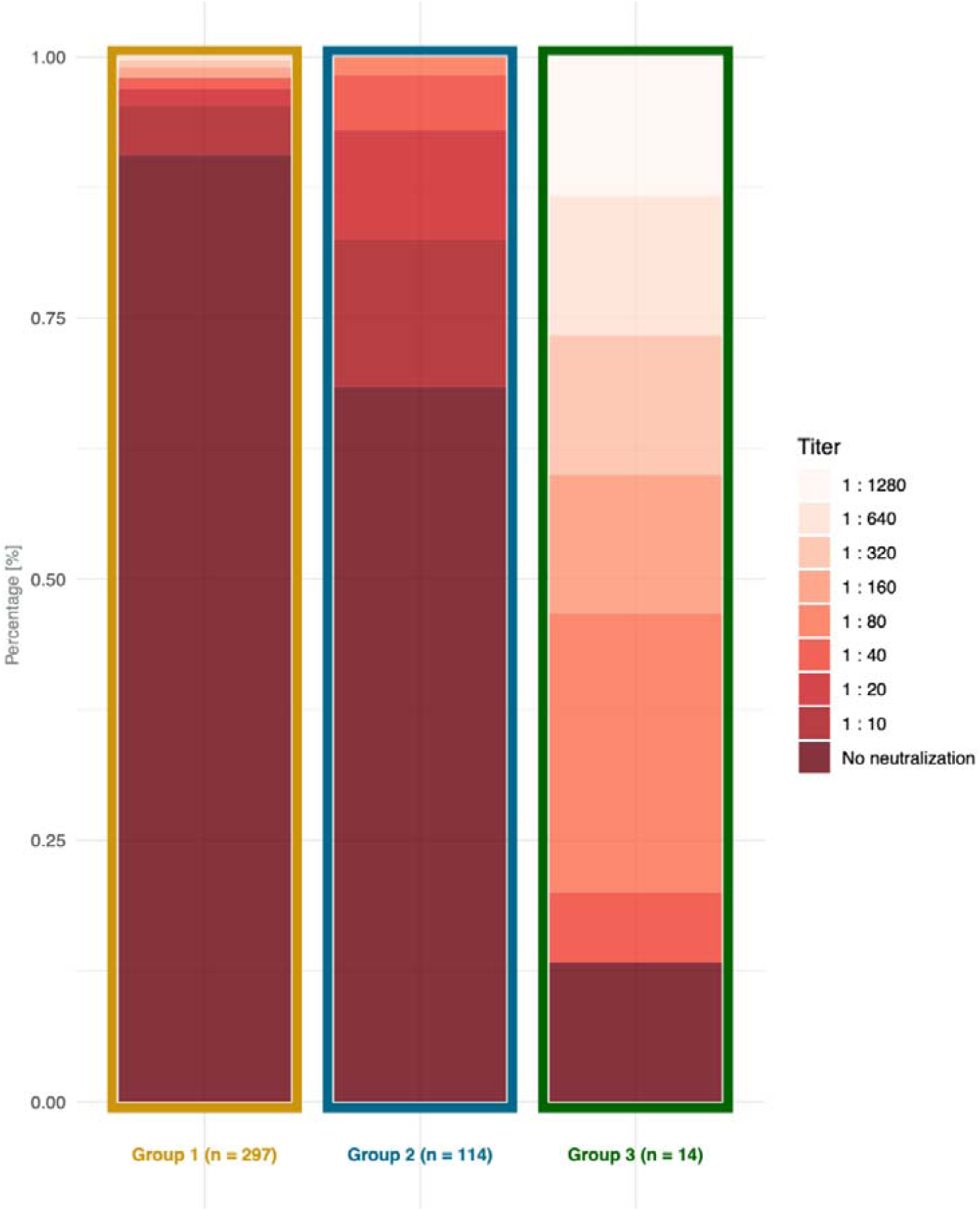
Barplots depicting the fraction of study participants in group 1 (residents of LTCFs ≥ 75 years of age), group 2 (HCWs at LTCFs, 18 to 70 years of age), and group 3 (residents of LTCFs after breakthrough infection) who exhibit neutralization titers against the Delta variant five to seven months after receiving the second dose of an mRNA vaccine.

A breakthrough infection vastly increased both the antibody concentration and fraction of individuals with detectable Delta-neutralizing antibodies among the elderly study participants, demonstrating recoverability of the humoral immune response.

These data support that a booster vaccine would be beneficial in this high-risk group of patients six months or earlier after completing the two-dose schedule with an mRNA vaccine. This is in coherence with other studies in this age group, even though studies based on pseudovirus neutralization assays provided divergently higher rates of neutralization.^17^ A possible explanation might be that for the neutralization assays we employed an authentic Delta variant isolate^16^ rather than a pseudovirus. Clinical isolates harbor additional mutations outside the Spike region which can impact the viral fitness or sensitivity to antibodies in cell culture. Markers of the cellular response of the study participants are currently examined and will be published when available. These additional data may influence the interpretation of the study results.

Since a majority of HCWs from these facilities, a group at large risk of transmitting infection to vulnerable patients, also did not have detectable neutralizing antibodies against the Delta variant, a booster of the humoral response appears indicated in this group as well.

Of the study participants in group 3, all of which had had a PCR-confirmed breakthrough infection, only 28.6% had detectable anti-nucleocapsid IgG response. This highlights that infection with SARS-CoV-2 in this group may occur without mounting an anti-nucleocapsid response, and some of the other participants may also have had an unknown infection that went undetected. Additional limitations of our study include that break-through infections in group 3 occurred at diverse time points before blood samples were taken. In addition to age, immunogenicity and efficacy of vaccines may be influenced and suppressed by several factors, including other medical conditions, and body composition with a very low or high BMI.^18^

Analyses of these factors in our study cohort will be provided in a subsequent publication.

## Data Availability

All data produced in the present work are contained in the manuscript

## Acknowledgments

The authors thank the staff of the facilities we visited, for their aid in conducting our investigation, and their tireless work in the pandemic and beyond.

We also thank all residents and staff who donated blood for this study.

